# Low frequency BOLD oscillations, *APOE4,* and plasma pTau_217_

**DOI:** 10.1101/2025.11.25.25340991

**Authors:** Trevor Lohman, Arunima Kapoor, Allison C Engstrom, Jillian Joyce, Melanie Quiring, John Paul M. Alitin, Aimee Gaubert, Amy Nguyen, Elizabeth Head, Rond Malhas, Kathleen E Rodgers, David Bradford, Basant Lashin, S. Duke Han, Mara Mather, Daniel A Nation

## Abstract

**Background:** Intrinsic low frequency oscillations in BOLD signal (BOLD-LFOs) are generally considered nuisance signal in connectivity analysis and discarded. However, recent evidence suggests BOLD-LFOs may shed light on cerebrovascular dysfunction and early Alzheimer’s disease pathophysiology, but the mechanisms remain unclear. No investigations to date have assessed the relationship between BOLD-LFOs and plasma pTau_217_, or how this relationship differs in apolipoprotein-e4 (*APOE4*) carriers who are vulnerable to cerebrovascular dysfunction and predisposed to AD pathophysiology.

**Methods:** Independently living older adults (N=118) without major neurological or psychiatric disorder were recruited from the community. Participants underwent resting-state brain functional MRI and venipuncture. Total BOLD-LFOs were quantified as signal power within the 0.01–0.10 Hz frequency range. Plasma level of pTau_217_ was assessed and linear regression was used to quantify the interactive effect of *APOE4* carrier status and BOLD-LFOs on plasma pTau_217_. 2×2 ANCOVA was used to compare BOLD-LFOs across *APOE4* carrier and amyloid positivity statuses based on previously reported pTau_217_ cutoffs.

**Results:** The interactive effect of *APOE4* carrier status and BOLD-LFO power was significantly associated with plasma pTau_217_ (β=−.65, p=.004). This relationship was driven by an inverse relationship between BOLD-LFOs and plasma pTau_217_ in *APOE4* carriers (β=−.49, p=.003). Amyloid-β (+) *APOE4* carriers displayed lower BOLD-LFOs than amyloid-β (−) *APOE4* carriers (p=.02) and amyloid-β (+) *APOE4* non-carriers (p=.04). All models were adjusted for age and sex.

**Conclusion:** Present study findings suggests that BOLD-LFOs are implicated early in AD pathophysiology in an *APOE4* dependent manner, adding support for the continued study of BOLD-LFOs in the context of cerebrovascular contributions to AD genetic risk.

## Background

Spontaneous blood-oxygen-level-dependent signal low-frequency oscillations (BOLD-LFOs) in the 0.01-0.1 Hz frequency range were once considered nuisance signal in fMRI connectivity analysis, caused by cardiac pulsations and respirations^1^. However, more recent work has demonstrated that other neurophysiological processes of potential relevance to brain health contribute to BOLD-LFOs, including spontaneous cerebrovascular reactivity^2,3^, astrocyte-mediated vasomotion^4–6^ and neuron-astrocyte crosstalk^7^. A growing body of evidence also indicates that there is disease-dependent variation in BOLD-LFOs^8,9^, suggesting changes in BOLD-LFOs may have relevance to the pathophysiology of some brain diseases and could hold unique diagnostic and clinical value relative to standard fMRI^9^.

For example, widespread dampening of the amplitude of BOLD-LFOs has been observed in Alzheimer’s Disease (AD) dementia patients^8^, where it correlates specifically with neuronal hypometabolism on ^18^F fluorodeoxyglucose-PET (FDG-PET). Another recent study found that BOLD-LFOs are altered early in the preclinical stages of AD, even in the absence of macrostructural atrophy^9^. This study also found that decreased amplitude of BOLD-LFOs correlates with the degree of diffuse cerebral amyloidosis and with accumulation of tau pathophysiological change in the entorhinal cortex^9^. Together these findings suggest that BOLD-LFO power attenuation occurs in the early stages of AD pathophysiology and is related to key amyloid, tau, and neurodegeneration markers of the disease. However, the molecular basis of these associations between BOLD-LFOs and AD remains unknown, and no studies to date have investigated how BOLD-LFOs relate to AD pathophysiology in the context of the AD risk gene, apolipoprotein-ε4 (*APOE4*). Finally, no studies have examined whether the amplitude of BOLD-LFOs are related to blood-based biomarkers of AD, including plasma pTau_217_.

The present study aims to address this knowledge gap by investigating the relationship between BOLD-LFOs and plasma pTau_217_, a reliable diagnostic and prognostic marker of AD progression^10^, in older *APOE4* carriers compared to non-carriers. The present investigation also sought to examine early changes in BOLD-LFOs by comparing BOLD-LFO power between *APOE4* carriers who have not yet developed pTau_217_ abnormality to *APOE4* carriers who have developed pTau_217_ based on previously established cutoffs for detecting cerebral amyloidosis^11^.

## Methods

### Participants

Participants were recruited from Los Angeles County and Orange County communities through outreach events, mailing lists, word-of-mouth, online portals, a research volunteer registry, and through the local Alzheimer’s Disease Research Center (ADRC). All procedures were conducted as part of the Vascular Senescence and Cognition (VaSC) Study at the University of Southern California (USC) and the University of California Irvine (UCI). Older adults aged 60 to 89 years who were living independently were included (**Table 1**). Study exclusions were a prior diagnosis of dementia, history of clinical stroke, family history of dominantly inherited neurodegenerative disorders, current neurological or major psychiatric disorders that may impact cognitive function, history of moderate-to-severe traumatic brain injury, active substance abuse, current use of medications impairing the central nervous system, current organ failure or other uncontrolled systemic illness, and contraindications for brain MRI. Eligibility for the study was verified by a structured clinical health interview and review of current medications with the participant and, when available, a knowledgeable informant study partner. This study was approved by the USC and UCI Institutional Review Board, and all participants gave informed consent. The anonymous data that support the findings of this study are available upon reasonable request from the corresponding author, DAN, through appropriate data sharing protocols.

**Table 1:** Participant characteristics and demographics grouped by *APOE4* carrier status.

| Variable Name | *APOE4* non-carriers (n=69) | *APOE4* Carriers (n=49) | *p* |
| --- | --- | --- | --- |
| Age (years) | 70.5±7.4 (60-89) | 69.2±6.7 (60-89) | .30<sup>a</sup> |
| Sex (n, female %) | 49 (71.0) | 31 (63.3) | .42<sup>b</sup> |
| BOLD-LFOs (0.01-0.1 Hz power) | 98.82±81.1 | 103.7±93.1 | .77<sup>a</sup> |
| Plasma pTau<sub>217</sub> (pg/ml) | .30±.17 | .42±.27 | .01<sup>a</sup> |
| Amyloid-β status<sup>c</sup> (n, amyloid+ %) | 12 (17.4) | 17 (34.7) | .04<sup>b</sup> |

***APOE4***: apolipoprotein e4 allele, **BOLD-LFO**: gray matter blood oxygen level dependent low frequency oscillations (0.01-0.1 Hz). a: independent t-test, b: chi-squared test. c: Amyloid-β status determined based on previously reported pTau<sub>217</sub> cutoff of .44 pg/ml which displays high combination sensitivity/specificity for detecting cerebral amyloidosis based on cerebrospinal fluid Aβ<sub>42/40</sub><sup>11</sup>.

## Measures

### Neuroimaging

All participants underwent brain MRI scans conducted on a 3T Siemens Prisma scanner with 20-channel head coil. High-resolution 3D T1-weighted anatomical (Scan parameters: TR□=□2300 ms; TE□=□2.98 ms; TI□=□900 ms; flip angle□=□9 deg; FOV□=□256 mm; resolution□=□1.0□×□1.0□×□1.2 mm^3^; Scan time□=□9 min) images were acquired, using 3-dimensional magnetization-prepared rapid gradient-echo (MPRAGE) sequences. Resting state fMRI scans comprised 140 contiguous echo-planar imaging (EPI) functional volumes (TR□=□3,000 ms, TE□=□30 ms, FA□=□80°, 3.3□×□3.3□×□3.3 mm voxels, matrix□=□64□×□64, FoV□=□212 mm, 48 slices).

Resting-state functional MRI data were preprocessed using a custom Python pipeline that integrates FMRIB Software Library (FSL v6.0), NiBabel, and SciPy. Preprocessing was conducted on each participant’s BOLD image series and corresponding T1-weighted anatomical scan.

Each participant’s T1-weighted anatomical image was skull-stripped using FSL’s BET and segmented into gray matter and white matter probability maps using the FAST tool. These maps were linearly registered to the participant’s functional space using FLIRT^12^, and binary tissue masks were generated by thresholding the probabilistic segmentations at 0.5. The fully preprocessed functional images were then registered to MNI152 standard space (2 mm resolution) using affine registration with 12 degrees of freedom.

The first 10 volumes of each BOLD time series were discarded. Slice timing correction was applied to adjust for inter-slice acquisition delays, as well as motion correction using FSL’s MCFLIRT tool to realign all volumes to reference image. After realignment, linear trends were removed from each voxel’s time series using a Python-based detrending procedure implemented with SciPy. The detrended data were spatially smoothed with a Gaussian kernel (4 mm full width at half maximum, approximated by sigma = 1.7 voxels) and temporally band-pass filtered to extract frequencies between 0.01-0.10 Hz using FSL’s fslmaths command.

Mean BOLD time series were extracted from the filtered functional images using the previously created gray matter mask. For each extracted time series, power spectral density (PSD) was calculated via fast Fourier transform, and total power within the 0.01–0.10 Hz frequency band was computed.

### Plasma AD Biomarkers

pTau_217_ concentrations were quantified in human plasma samples using the ultra-sensitive Simoa (Single Molecule Array) assay platform developed by Quanterix (Lexington, MA, USA). The pTau_217_ assay utilized a bead-based sandwich 3 step immunoassay format run on the Quanterix HD-X Analyzer, which allows for digital quantification of low-abundance biomarkers in biological fluids. Plasma samples were collected in K2EDTA tubes, centrifuged at 2000 x g for 10 minutes at 4°C, and stored at −80°C until analysis. Prior to assay, samples were thawed on ice and centrifuged again to remove any debris. The pTau-217 assay was performed according to the manufacturer’s instructions (Quanterix pTau-217 Advantage Kit, Catalog # 104588), using 100 μL of plasma per well. Capture antibodies specific for pTau-217 were immobilized on paramagnetic beads, while detector antibodies were labeled with a proprietary enzyme tag. Following incubation and washing steps, individual immunocomplexes were transferred to a microwell array, allowing for single-molecule detection. A chemiluminescent substrate was added, and signals were captured digitally by the HD-X Analyzer.

A plasma pTau_217_ cut off of 0.44 pg/ml has previously displayed high combination specificity and sensitivity (>85%) for detecting cerebral amyloidosis based on cerebrospinal fluid Aβ42/40^11^.This cutoff was used in the present study to categorize participants as either amyloid-β positive or negative.

### *APOE* Genotyping

Fasted blood samples were obtained by venipuncture and used to determine participant *APOE* genotype. Genomic DNA was extracted using the PureLink Genomic DNA Mini Kit (Thermo). *APOE* genotyping was performed as previously described^13–15^. *APOE4* carriers were defined as participants with at least one copy of the apolipoprotein-ε4 allele. All analyses were performed at the same lab at the University of Arizona (KR).

### Statistical Analyses

121 participants underwent brain fMRI, anatomical T1w MRI, and venipuncture. All data were screened for outliers (±3 SD) and three gray matter BOLD-LFO outliers were identified and removed with values of +4.84 SD, +4.22 SD, and +3.79 SD. After outlier screen, the total analyzed sample was N=118. All assessed variables were compared between *APOE4* carriers and non-carriers using independent t-tests.

Data analyses were performed in R. Hayes PROCESS Model 1 (simple moderation) was used to assess the potential moderating effect of *APOE4* carrier status on the relationship between BOLD-LFOs and plasma pTau_217_, where x=BOLD-LFO, Y=pTau_217_, and w=*APOE4* carrier status. This analysis was repeated excluding participants with a copy of the *APOE2* allele (n=4). Linear regression assumptions regarding linearity, multicollinearity (VIF<5), and homoscedasticity (Breusch-Pagan test) were met.

Lastly, within group analyses were performed to assess the relationship between BOLD-LFOs and pTau_217_ within the *APOE4* carrier and non-carrier groups. All models were adjusted for age and sex. 2X2 ANCOVA was used to compare BOLD-LFOs by amyloid-β positivity and *APOE4* carrier status adjusted for age and sex, and pairwise comparisons were also performed. For illustration purposes, power spectral density plots were generated to compare BOLD-LFOs power between amyloid-β (+) and amyloid-β (−) *APOE4* carriers.

Minimum detectable effect sizes given a power of .80, alpha of .05, two covariates, and a sample size of N=118 was calculated using the pwr package in R as .4R^2^=.05 for the BOLD-LFO\**APOE4* interaction effect. False discovery rate correction was used to account for multiple comparisons in the primary analysis including *APOE4*\*BOLD-LFOs interaction effect and subgroup analyses (within *APOE4* carrier and non-carrier group effects)^16^.

## Results

118 participants were included for analysis, participant characteristics and demographics for this sample are shown in **Table 1** grouped by *APOE4* carrier status. BOLD-LFOs summary z-map displayed in **Figure 1**.

**Figure 1:**
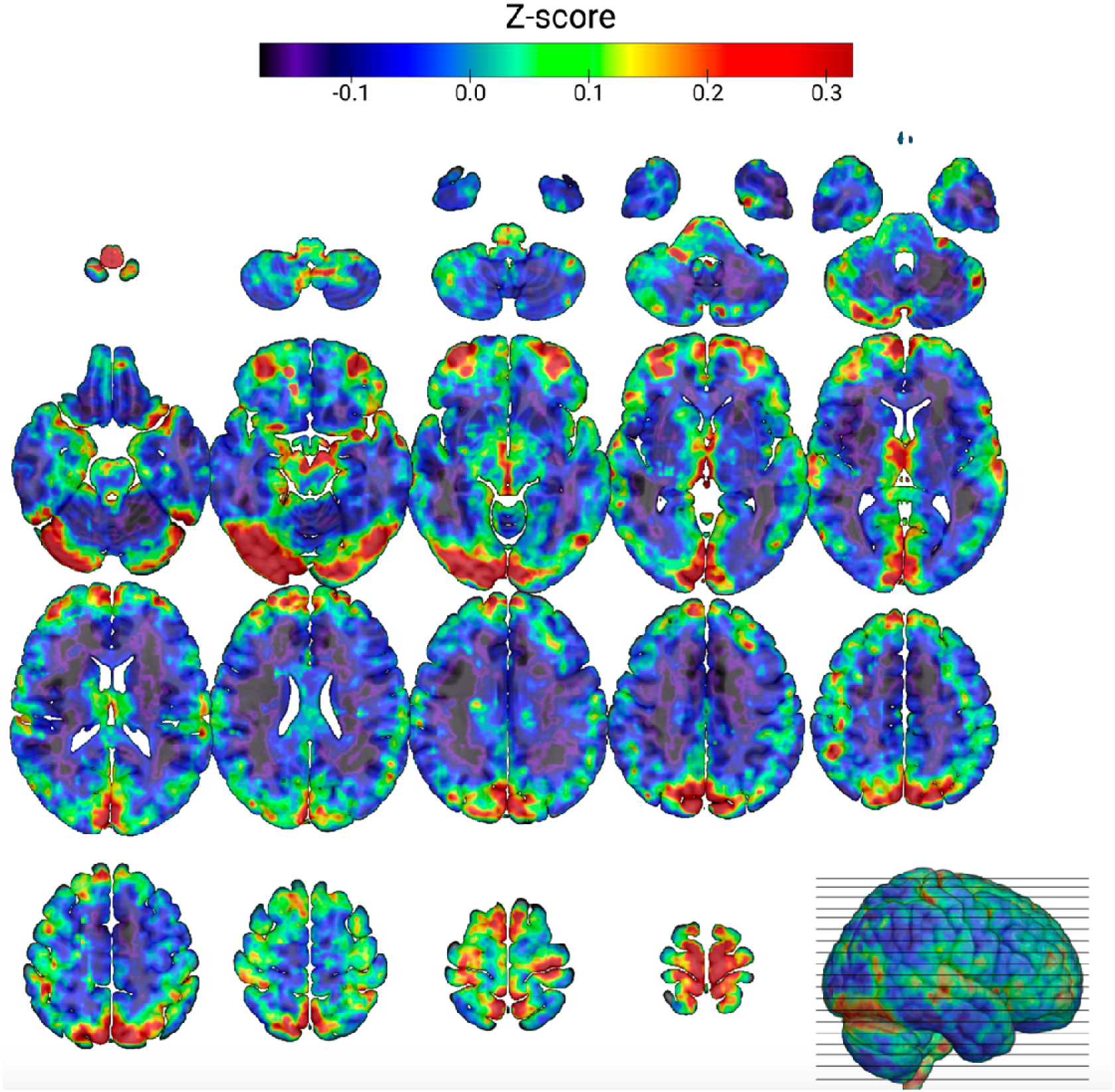
0.01-0.1Hz BOLD low frequency oscillations (BOLD-LFO) summary Z-map (N=118). BOLD-LFO power z-map registered to the MNI152 2 mm template. Individual subject z-maps were computed by band-limited Fourier analysis of preprocessed BOLD time series, excluding CSF voxels and non-brain regions. The resulting subject maps were normalized within brain masks and averaged across the cohort. Warmer colors indicate higher relative low-frequency power compared to the whole-brain mean.

The interactive effect of *APOE4* carrier status and gray matter BOLD-LFOs was significantly associated with plasma pTau_217_ (β=−.65, p=.004). This was driven by an inverse relationship between BOLD-LFOs and pTau_217_ in *APOE4* carriers (β=−.49, p=.003) as shown in **Figure 2**. This analysis was repeated excluding participants with a copy of the *APOE2* allele (n=4). The interaction effect was similar for this analysis (β=−66 p=.01), and when adjusted for age and sex (β=−.67 p=.007). Additional sensitivity analyses were performed for all tests adjusting for default mode network connectivity which did not attenuate the observed significant relationships. The BOLD-LFO\**APOE4* interaction term’s relationship with pTau_217_, and subsequent within group analysis results survived correction for multiple comparisons.

**Figure 2:**
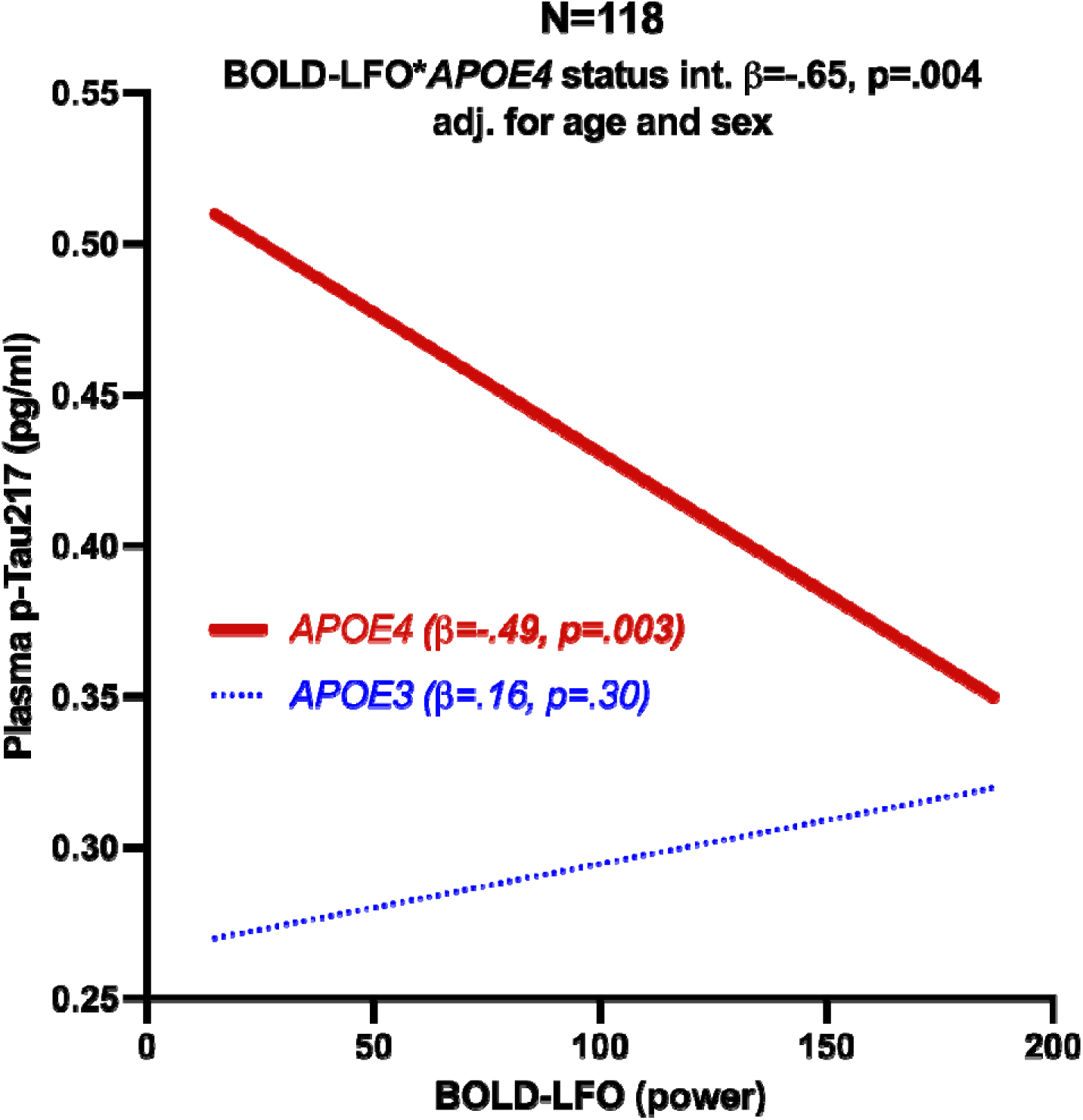
The relationship between 0.01-0.1Hz gray matter BOLD low frequency oscillations (BOLD-LFO) and pTau_217_ in *APOE4* carriers (n=49) compared to *APOE3* homozygotes (n=69). The effect of BOLD-LFOs on pTau_217_ conditional upon *APOE4* carrier status (interaction effect) is reported as standardized regression coefficient (β) and p-value in the total sample(N=118). Within group statistics reported in figure legend as β and p-value within *APOE4* carrier status groups.

To visualize this effect across pTau_217_ cutoffs and *APOE4* carrier status groups, a 2X2 ANCOVA with pairwise comparisons was performed. The amyloid-β positivity status\**APOE4* carrier status interaction was statistically significant (p=.006). In pairwise comparisons amyloid-β positive *APOE4* carriers displayed lower BOLD-LFO power than amyloid-β positive *APOE3* homozygotes (p=.04), and amyloid-β negative *APOE4* carriers (p=.02) (**Figure 3**).

**Figure 3:**
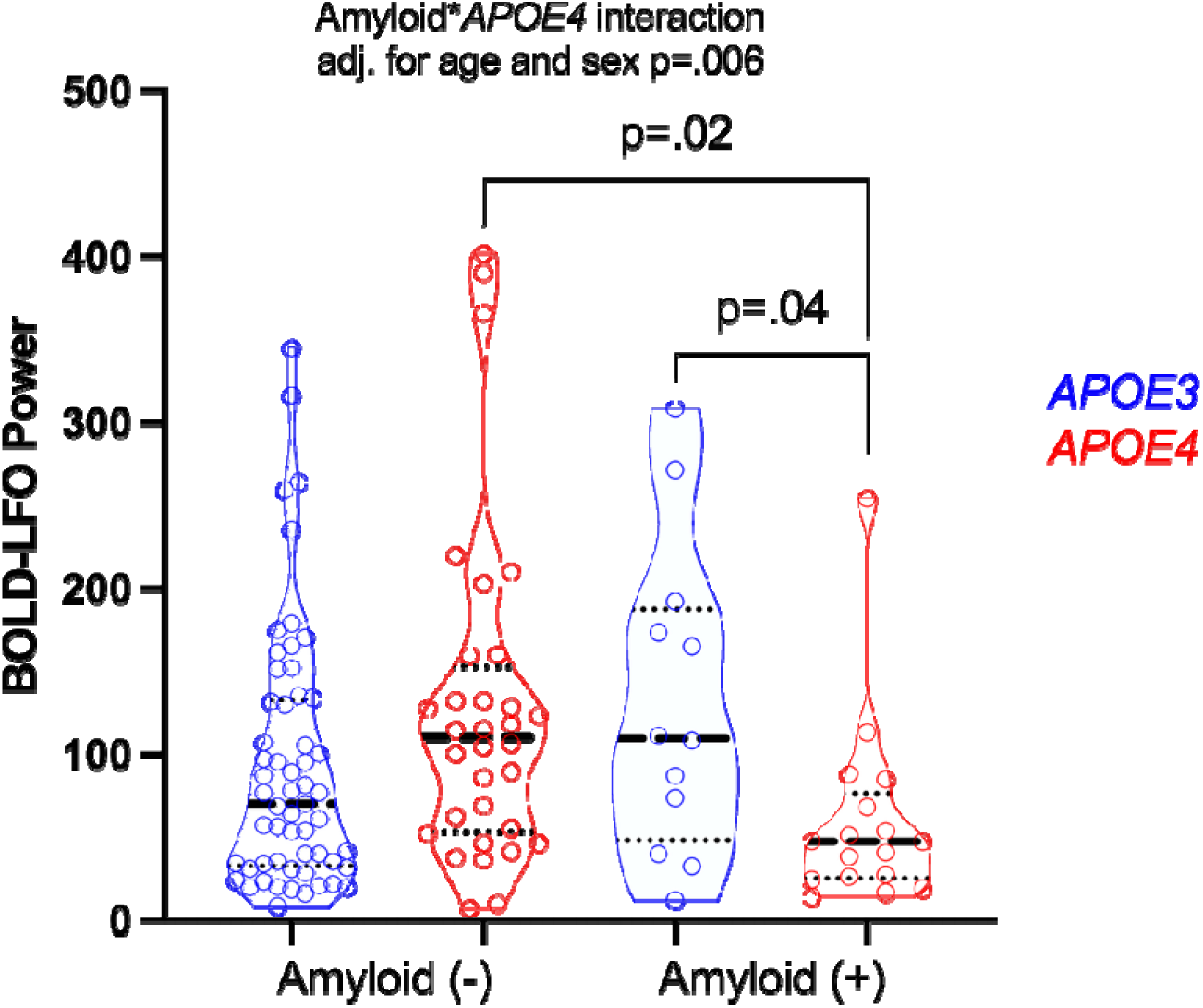
Gray matter blood oxygen level dependent low frequency 0.01-0.10 Hz oscillations (BOLD-LFO) comparisons of *APOE4* status (*APOE3* homozygotes vs. *APOE4* carriers) by amyloid-β positivity status. 2X2 ANCOVA interaction model and subsequent pairwise comparisons are adjusted for age and sex. Amyloid-β status determined based on previously established pTau_217_ cutoff of .44 pg/ml which displays high combination sensitivity/specificity for detecting cerebral amyloidosis based on cerebrospinal fluid Aβ42/40^11^.

A power spectral density plot is shown in **Figure 4** to visualize the BOLD-LFO differences between amyloid-β positive and negative *APOE4* carriers across the full spectrum of low frequency oscillations.

**Figure 4:**
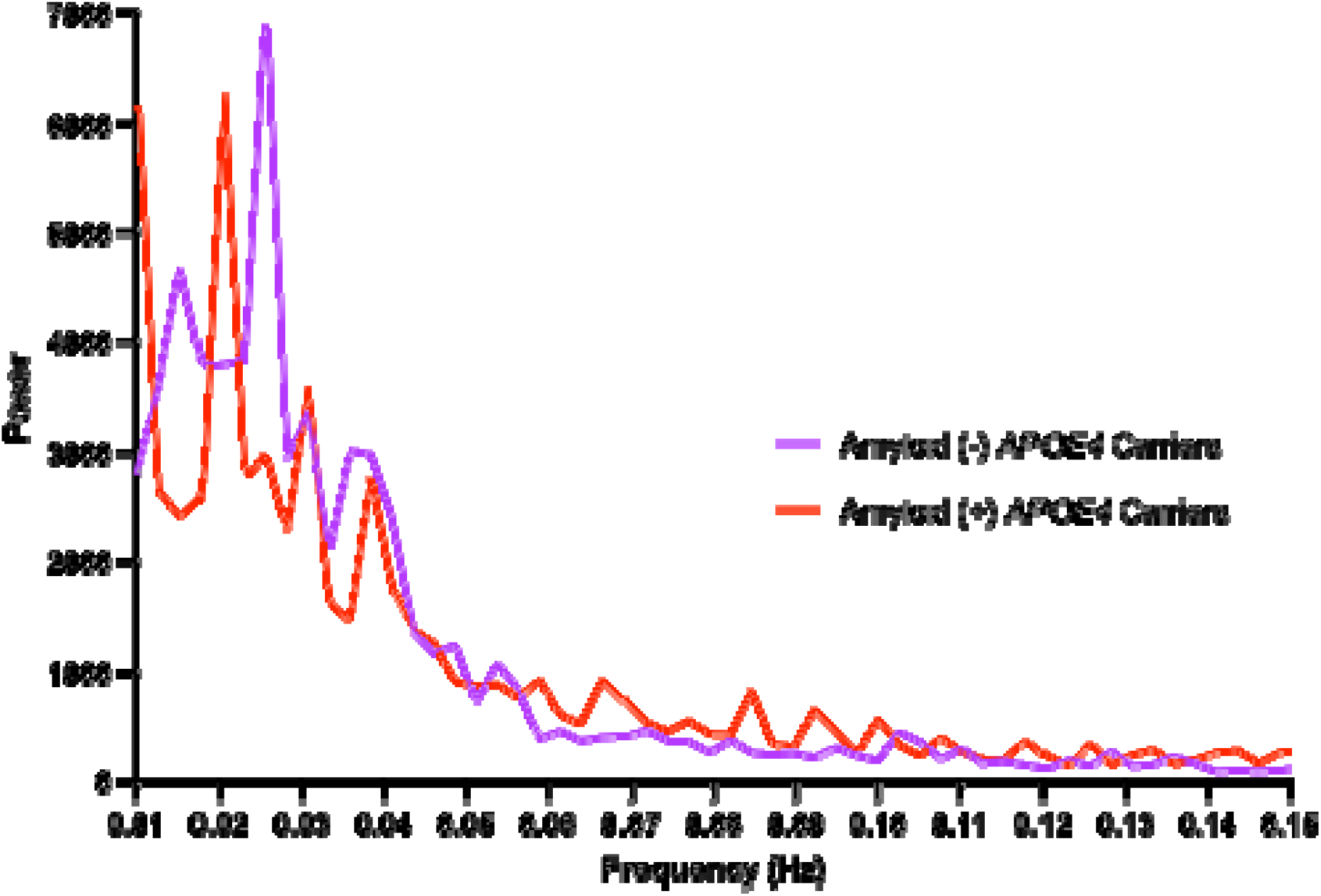
Gray matter blood oxygen level dependent low frequency oscillations (BOLD-LFO) power spectral density plot. Comparisons across amyloid-β positivity status (amyloid-β positive= pTau_217_>.44 pg/ml) in APOE4 carriers. Amyloid-β positive n=17, amyloid-β negative n=32. Amyloid-β status determined based on previously reported pTau217 cutoff of .44 pg/ml which displays high combination sensitivity/specificity for detecting cerebral amyloidosis based on cerebrospinal fluid Aβ42/40^11^.

## Discussion

The present study finds that BOLD-LFO power is inversely related to plasma pTau_217_ levels in *APOE4* carriers but not in non-carriers, suggesting a role for the *APOE4* gene in the association between BOLD-LFOs and AD pathophysiological change. Findings also suggest this alteration in BOLD-LFOs may be observed in the very earliest stages of AD pathophysiological change, as evidenced by decreased BOLD-LFO power in amyloid-β positive *APOE4* carriers relative to amyloid-β negative *APOE4* carriers. Together these results are consistent with prior work suggesting changes in BOLD-LFOs are related to early AD pathophysiological changes on PET and CSF biomarkers and extend prior findings by demonstrating the relationship between decreased BOLD-LFOs and early stage plasma pTau_217_ in *APOE4* carriers specifically.

*APOE4* is associated with vascular, neuroinflammatory, and metabolic changes in neurons^17,18^ and neuroglia^19–22^. One or more of these neurophysiological changes could be implicated in the relationship between BOLD-LFOs and pTau_217_ in *APOE4* carriers specifically. For example, *APOE4* conveys susceptibility to cerebrovascular dysfunction^23–25^, which could contribute to the observed decreased in BOLD-LFOs through decreased pulsatile or intrinsic vasomotion^2,3^. Astrocytes are also increasingly thought to be important contributors to BOLD-LFO signal^7,26^ and *APOE4* is associated with numerous changes to astrocyte function caused by an accumulation of lipid droplets and a buildup of unsaturated fatty acids within astrocytes^19^. This accumulation of lipid droplets is sufficient to induce astrocyte reactivity, triggering the secretion of inflammatory chemokines and cytokines^27,28^.

An examination of BOLD-LFO differences in amyloid-β positive and amyloid-β negative *APOE4* carriers in the present study suggests that the largest differences in power exist in the lower frequency ranges, which are associated with astrocyte-mediated vasomotion^4,5,7,26,29^, particularly vasodilation^6^. Thus, the observed relationship between reduced BOLD-LFOs and AD pathophysiology in APOE4 carriers may be related to changes in vascular function, astrocyte reactivity, or astrocyte-vascular interactions^30^. However, other systemic physiological processes like cardiac pulsations and respiratory dynamics contribute to BOLD-LFO signal, and further studies are needed to determine the specific mechanistic contributions to BOLD-LFOs that may be implicated in *APOE4* associated AD pathophysiology.

Strengths of the present study include the study of BOLD-LFOs in early AD pathophysiological change by comparing *APOE4* carriers to non-carriers with and without pTau_217_ abnormality. Limitations include the cross-sectional nature of the study and the lack of a universally accepted plasma pTau_217_ cutoff for determining cerebral amyloidosis. The present study findings suggest there should be further investigation into the potential role of decreased amplitude of BOLD-LFOs in *APOE4*-related AD pathophysiological changes with potential implications for our understanding of early-stage AD and related diagnostic approaches.

## Data Availability

The anonymous data that support the findings of this study are available upon reasonable request from the corresponding author, DAN, through appropriate data sharing protocols.

